# Something from nothing: Sensitivity and specificity of Xpert MTB/RIF Ultra on contaminated liquid cultures for tuberculosis and rifampicin-resistance detection

**DOI:** 10.1101/2022.12.07.22283223

**Authors:** YT Ghebrekristos, N Beylis, CM Centner, R Venter, B Derendinger, H Tshivhula, S Naidoo, R Alberts, B Prins, A Tokota, T Dolby, FM Marx, SV Omar, R Warren, G Theron

## Abstract

**Background:** Xpert MTB/RIF Ultra (Ultra) (Cepheid, Sunnyvale, USA) is a widely-used rapid front-line TB and rifampicin susceptibility test. Mycobacterium Growth Indicator Tube 960 (MGIT960) culture is still used as an adjunct for tuberculosis (TB) and drug susceptibility diagnosis but is vulnerable to contamination. Whether Ultra can be used on to-be-discarded contaminated cultures is uninvestigated.

**Methods:** We stored contaminated MGIT960 tubes (growth-positive, acid-fast-bacilli-negative) inoculated to diagnose pulmonary TB in a routine high-volume laboratory in Cape Town, South Africa. Patients who had, at contamination-detection, no positive TB results (smear, Ultra, culture) and another specimen submitted three months post-contaminated specimen submission were selected. We evaluated the sensitivity and specificity of Ultra on contaminated growth from the first culture for 1) TB (next-available non-contaminated culture result reference standard), and 2) rifampicin resistance (vs. MTBDR*plus* on the later isolate). We calculated potential time-to-diagnosis improvements. We also evaluated MPT64 TBc (TBc).

**Findings:** 2186 patients had a contaminated diagnostic culture. 49% (1068/2186) had no other specimen submitted, despite guidance to the contrary. After 319 ineligible patients were excluded, 799 with at least one repeat specimen submitted remained: 31% (n=246), 54% (n=429) and 16% (n=124) were repeat-specimen culture-positive, -negative, and -contaminated, respectively. When Ultra was done on the initial contaminated growth, sensitivity and specificity were 89% (95% CI 84-94) and 95% (90-98) for TB and 95% (75-100) and 98% (93-100) for rifampicin-resistance. If our approach were performed the day after initial contamination detection, time-to-TB-detection would improve a median (IQR) of 23 (13-45) days and, importantly, provide a result in many patients who had none. TBc had poor accuracy.

**Conclusion:** Ultra on acid-fast-negative growth from contaminated MGIT960 tubes had high sensitivity and specificity; approximating World Health Organization-target product performance sputum test and exceeding drug susceptibility testing (DST) criteria. Our approach could mitigate contamination’s negative effects, especially when repeat specimens are not submitted.

**Research in context:** *Evidence before this study:* Improving the diagnosis of tuberculosis (TB) and drug-resistance through strengthening the laboratory care cascade is a public health priority. Scale-up of molecular tests like Xpert MTB/RIF Ultra (Ultra) (Cepheid, Sunnyvale, USA), for the upfront diagnosis of TB and rifampicin-resistance has doubtlessly improved the care cascade, however, culture, despite several limitations, continues to be used for the diagnosis and susceptibility testing for technical, historic, and cost reasons (the most common TB culture platform is the MGIT960 liquid culture system). The fact that global TB diagnosis still, in part, relies on culture means that culture-contamination, which represents a failed attempt at testing, worsens care cascade gaps. Contamination requires another specimen to be collected from patients, however, this causes delays or complete care cascade drop out of patients. Contaminated cultures are traditionally checked with microscopy to see if they contain acid-fast bacilli (AFB), however, the use of Ultra on contaminated cultures, especially those who are AFB-negative, is unexplored. If performance is high, the negative impact of culture-contamination, which is frequent in many settings, could be drastically mitigated as Ultra is widely-available.

*Added value of this study:* We showed that Ultra on to-be discarded contaminated MGIT960 cultures can detect TB in a highly sensitive and specific manner (89% sensitivity, 95% specificity). It also had excellent sensitivity and specificity for rifampicin resistance (95% sensitivity, 98% specificity). Performance levels exceeded those accepted by the World Health Organization for Ultra done directly on respiratory specimens. In patients who, after initial culture contamination had another specimen submitted for culture, our approach could reduce time to diagnosis by approximately 23 days. Critically, many patients with contamination had, despite programmatic guidance, no record of a further attempt to diagnose TB (44%), and in these patients our Ultra on contaminated cultures approach would result in an accurate TB and rifampicin-resistance result where none would ordinarily occur.

*Implications of all available evidence:* When done on contaminated MGIT960 culture growth resulting from a failed attempt to diagnose TB, Ultra has excellent performance for TB and rifampicin-resistance detection and would likely reduce the impact of culture-contamination on the diagnostic care cascade. Laboratories should consider evaluating and potentially implementing this approach wherever TB culture is done for diagnostic purposes.

## Introduction

Rapid molecular tests are essential in the fight against tuberculosis (TB). The World Health Organization (WHO) endorsed Xpert MTB/RIF Ultra (Ultra) (Cepheid, Sunnyvale, USA) as the initial test for all patients with signs and symptoms of TB due to Ultra’s short turn-around-time (TAT) and low *Mycobacterium tuberculosis* complex (MTBC) limit of detection (1-3). Importantly, as with most PCR tests, targeted MTBC DNA amplification and detection can occur in the presence of contaminating DNA.

Despite limitations, mycobacterial culture is often performed for initial TB diagnosis. Reasons are multifactorial and setting-dependent but include clinically-justified scenarios that involve presumptive TB patients with a negative Ultra and HIV (and/or clinical worsening) (4), symptomatic patients with recent previous TB (5) (residual DNA from previous TB episode causes false-positive Ultra results (6)), special groups like children and practical reasons e.g. if GeneXpert capacity is diminished. Culture is also routinely used isolate bacilli for drug susceptibility testing (DST) (7, 8). The Mycobacterium Growth Indicator Tube 960 system (MGIT960; Becton Dickinson Diagnostic Systems, Sparks, USA) is the preferred culture method due to sensitivity and automatability, however, increased contamination rates can be a major drawback (9).

Prior to MGIT960 culture, specimens are decontaminated, centrifuged, and resuspended in buffer; an aliquot of which is used for inoculation. Decontamination differentially reduces the culturability of bacteria (mycobacteria are typically least affected), making contaminating overgrowth less likely. MGIT960 growth is automatically monitored, and after a tube is flagged as growth-positive, an acid-fast stain of growth is done. If acid-fast bacilli (AFBs) are observed (irrespective of other cells’ presence), an antigen or PCR test is done to confirm MTBC. If growth occurs but no AFBs are observed that specimen is reported as culture-contaminated and the specimen and derivatives discarded. An “acceptable” MGIT960 contamination rate is 3-8% according to the manufacturer (10), however, high contamination rates, often attributable to too low NaOH concentrations, are often described: 30% in Zambia (11), 24% in Burkina Faso (12), 17% in South Africa (9) and 15% in Ethiopia (13). After contamination, laboratories typically issue a request to health workers to re-submit a new specimen for repeat investigation. This consumes resources, creates a care cascade gap (a repeat specimen from patients may never be submitted), and delays diagnoses, including for drug-resistance.

The impact of MGIT960 contamination on initial diagnosis might be mitigated if AFB-negative growth did not signify the end of a specimen’s journey. Ultra is a logical test to evaluate; it has well-established superior sensitivity vs. smear on respiratory specimens, determines rifampicin susceptibility, is largely automated and, while scaled-up in many settings, is often underutilised (14), meaning spare capacity may exist. We evaluated the sensitivity and specificity and potential effect of Ultra applied to contaminated MGIT960 growth for the detection of TB and rifampicin susceptibility.

## Materials and methods

### Routine specimen processing and programmatic reporting of MGIT960 cultures

Specimens were processed in a high-throughput National Health Laboratory Service (NHLS) laboratory in Cape Town, South Africa using the standard NALC-NaOH procedure (1.0% final concentration) (10). Per internal standard operating procedures, 0.5 ml of the re-suspension were inoculated into a MGIT960 tube supplemented with PANTA (10) and incubated for a maximun of 35 days (standard in our programmatic laboratory due to limited incubator space). After a tube was flagged as growth-positive (200 growth units), Ziehl-Neelsen (ZN) microscopy was done to detect AFBs. TB identification from AFB-positive growth was done by line probe assay MTBDR*plus* (Hain Lifescience, Nehren, Germany), (if DST also required) or MPT64 TBc (TBc, Becton Dickinson, Sparks, USA) (if positive by either test, growth was reported as “culture-positive”). If only non-AFBs were observed the cultures are reported per programmatic policy as “*Culture contaminated with no further result to follow*”.

### Setting and contaminated culture selection

Contaminated cultures with no AFB observed from an attempt at an initial culture-based diagnosis on respiratory specimens (expectorated or induced sputum, tracheal aspirates) were consecutively collected in a three-month window (1 June 2019-31 August 2019) and stored at 2-8°C. Results of routine TB investigations (Ultra, MGIT960, MTBDR*plus*, TBc) on specimens or isolates up to three months after initial contamination detection were extracted (e.g., up until 30 November 2019 for contamination detected 31 August 2019; designated as the follow-up period) (**Figure 1**). For inclusion in diagnostic accuracy analyses, patients had to have at least one positive or negative culture result from later specimen(s) submitted within three months of the initial culture contamination report. To avoid bias, we did not preferentially select patients based on results from a later specimen (e.g., Ultra-positive on the repeat specimen) other than culture. We excluded patients with a known smear-, Ultra-, or culture-positive result before the initial contamination report (including those 12 months prior to contaminated specimen collection, which were deemed to be submitted for treatment monitoring), those who had no later culture-positive or -negative results (which lacked reference standard information for our primary analysis), and we ignored culture result(s) from any specimens submitted either on the same day as the specimen later found to be contaminated or while that initial specimen was still being incubated (in other words, we only included repeat culture results when the specimen for that repeat was submitted after issue of the initial contamination report; multiple simultaneous diagnostic cultures were non-compliant with the programmatic algorithm). If patients had more than one contaminated culture, the earliest was selected and only one contaminated culture per patient underwent Ultra or TBc.

**Figure 1.**
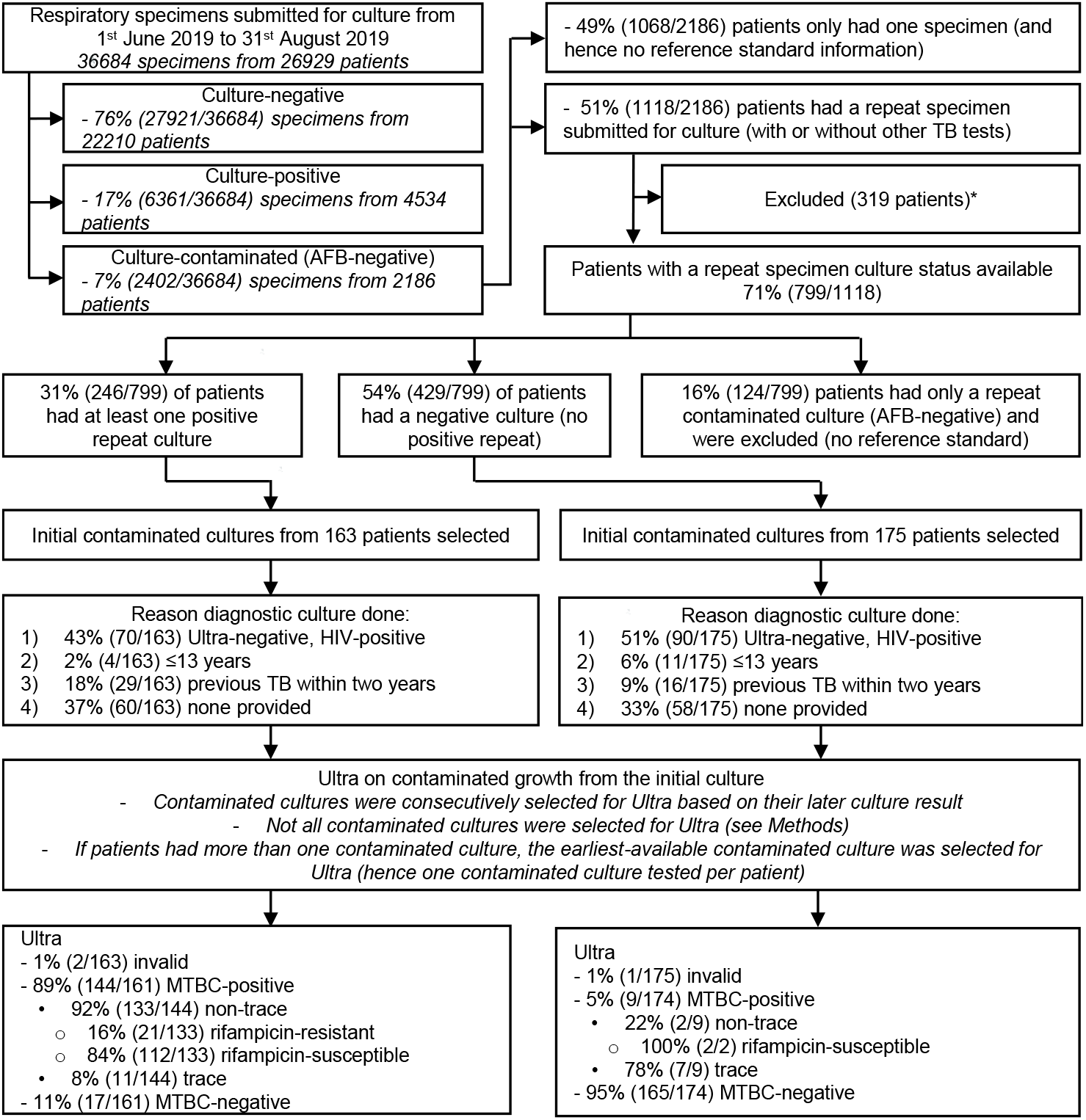
Study profile. We quantified the number of cultures done for TB diagnosis on respiratory specimens over a three-month period in a high-volume programmatic laboratory. 2402 specimens submitted for culture were AFB-negative culture-contaminated, 56% of which had a repeat specimen submitted for another culture within three months. After exclusion of repeat specimens submitted on the same day as the contaminated specimen, the repeat culture results of the remaining 1012 were captured. The initial contaminated growth tube of those who were repeat culture positive or negative was then retrieved from the facility’s storage and a consecutive subset selected for Ultra. For this subset, we retrieved the reason provided for the initial diagnostic culture request. Ultra on contaminated cultures had very low invalid rates, correctly detected 89% of reference standard-positive patients as having TB (and identified resistance in 14%) and detected 95% of reference standard-negative patients as Ultra-negative. Therefore, when done on AFB-negative contaminated growth, Ultra results are highly concordant with culture results from later submitted specimens, as well as a MTBDR*plus* result done from that later culture isolate. Abbreviations: AFB, acid-fast bacilli; MTBC, *Mycobacterium tuberculosis* complex; TB, tuberculosis; Ultra; GeneXpert MTB/RIF Ultra. *Repeat specimen submitted at the same time or while the culture-contaminated specimen was undergoing incubation and there was no later specimen submitted (n=193) or, even if there was a later specimen submitted, a culture-positive result occurred before the contamination report of the initial specimen was issued (n=126). In other words, we excluded specimens from patients who lacked reference information or in whom culture contamination would not have likely been consequential for patient management as multiple specimens for culture were submitted in a short period.

### Ultra and TBc testing of contaminated MGIT960 cultures

After eligible contaminated culture, cultures were separated based on their later culture result, a subset of 338 (163 later-culture-positives, 175 later-culture-negatives) were consecutively selected and processed for Ultra. 6 ml contaminated growth was centrifuged (3000*g*, 15 min) and supernatant discarded, leaving a ∼0.7 ml pellet resuspended by gentle mixing in 1.4 ml sample reagent (Cepheid, USA). The mixture was subsequently tested as recommended (15) (laboratory standard operating procedure in **Supplementary Material 1**). We did not include additional contaminated cultures because our sensitivity and specificity estimates were sufficiently precise (≤5% on either side of the confidence interval) (16). For TBc, contaminated growth that tested Ultra-positive (n=20) or Ultra-negative (n=20) were consecutively selected and tested as recommended (17) using 100 µl of contaminated growth.

### Reference standards

If at least one subsequent culture was MTBC-positive, the patient was classified as definite TB. If there was no MTBC-confirmed growth and no other MTBC-positive cultures from the follow-up period, the patient was classified as not TB. For rifampicin susceptibility, reference standard resistant cases were definite TB and MTBDR*plus* rifampicin-resistant on a subsequent isolate (reference standard susceptible cases hence were definite TB and MTBDR*plus* rifampicin-susceptible).

### Diagnostic accuracy and predictive value analyses

Sensitivity and specificity for TB and rifampicin susceptibility were determined using 2×2 tables with 95% confidence intervals (exact binomial method). We compared Ultra results between patients with previous TB (confirmed on a specimen submitted less than four years but more than one year before the contaminated specimen) to those with no previous TB to evaluate if, as observed with sputum (6, 18), residual DNA from previous episodes caused false-positive Ultra results. We calculated how PPV and NPV change as the frequency of the condition of interest (“prevalence”) changed in people who had another specimen submitted. For example, TB “prevalence” was defined as the proportion culture-positive in those patients who had 1) another specimen submitted within three months of the first contaminated culture and 2) that later specimen was culture-positive or -negative. For rifampicin-resistance, prevalence was defined as the proportion of patients who had another specimen submitted within three months-that was culture-positive and in whom the resulting isolate was successfully tested with MTBDR*plus*-who were MTBDR*plus* rifampicin-resistant on contaminated culture.

### Potential improvements in lost-to-follow-up, time-to-diagnosis and turn-around-time

We designated patients with a contaminated culture who had no record of any repeat specimen in the follow-up period as lost-to-follow-up. Diagnostic delay caused by contamination was defined as days between report of the initial contamination result and, if not lost-to-follow-up the earliest next-positive result (Ultra, smear, culture) on a later repeat specimen. If patients had repeat specimen results and none were positive, the earliest culture-negative result date was used. The difference in the initial specimen culture contamination report date and the repeat specimen culture result date was calculated (potential improved TAT).

### Ethics

This study received approval from the Human Research Ethics Committee Division of Molecular and Human Genetics, Department of Biomedical Sciences at Stellenbosch University (S20/08/189) and the NHLS Academic Affairs, Research and Quality Assurance (AARMS; PR2119347). As we used programmatically-submitted deidentified remnant material that would be discarded the need for written informed consent was waived.

## Results

### Quantity of routine diagnostic cultures

Between 1 June 2019 and 31 August 2019, 36684 specimens from 26929 unique patients were processed for diagnostic culture. 70% (18936/26929) patients had one specimen submitted for culture, accounting for 52% (18936/36384) of total specimens. The remaining 30% (7993/26929) patients had ≥2 specimens submitted, amounting to 48% (17748/36684) of overall specimen quantity.

### Initial culture results

76% (27921/36684) specimens were culture-negative, 17% (6361/36684) culture-positive, and 7% (2402/36684) culture-contaminated (without AFBs) (from 22210, 4534, and 2186 unique patients, respectively).

### Repeat specimen availability in initially culture-contaminated patients

49% (1068/2186) of patients with a culture-contaminated specimen had no further specimens submitted, representing care cascade loss (**Figure 1**). Of the 51% (1118/2186) of patients with at least one repeat specimen, 29% (319/1118) were excluded. Amongst the remaining 799 patients, 31% (246/799) had at least one repeat specimen culture-positive, 54% (429/799) had all repeat specimen(s) culture-negative (hence 675 eligible patients with reference standard information), and 16% (124/799) had only culture-contaminated results from repeat specimen(s). The median (IQR) days between first culture contamination report and the second specimen culture report date were 42 (30-54) overall.

### Xpert MTB/RIF Ultra on contaminated cultures

#### Results

163 culture-positive patients and 175 culture-negative patients were selected for Ultra testing. 1% 3/338 gave an Ultra non-actionable result (all invalid). For culture-positives, Ultra detected TB in 89% (144/161) of contaminated cultures [7% (10/144), 8% (12/144), 26% (37/144), 27% (39/144), and 32% (46/144) in the “trace”, “very low”, “low”, “medium”, “high” Ultra semi-quantitation categories, respectively]. Of the non-trace categories, 16% (21/133) were Ultra rifampicin-resistant. Of the culture-negatives, Ultra detected TB in 5% (9/174) of contaminated cultures (seven trace, two non-trace and both rifampicin susceptible).

#### Sensitivity and specificity

For TB, sensitivity and specificity was 89% (95% CI 84-94; 144/161) and 95% (90-98; 165/174), respectively. For rifampicin-resistance, sensitivity and specificity were 95% (75-100; 19/20) and 98% (93-100; 100/102), respectively.

#### “Trace” recategorization

If Ultra “trace” calls were excluded, sensitivity for TB remained 89% (83-93; 134/151) and specificity improved to 98% (96-100; 165/167), however, 5% (17/335) of patients were Ultra trace-positive on contaminated growth and would be excluded. If “trace” calls were reclassified to TB-negative, sensitivity decreased to 83% (77-89; 134/161) and specificity improved to 99% (96-100; 172/174).

#### Previous TB

Previous TB was more frequent in false-than true-positives [67% (6/9) vs. 17% (24/144); p=0.0001] and hence specificity reduced in patients with previous TB [67% (12/18) vs. 98% (153/156) in those with no previous TB; p<0.0001]. This did not change with different “trace” recategorization strategies (**Supplement**).

### Predictive value of Ultra on contaminated cultures for the diagnosis of TB and rifampicin resistance

#### TB

“Prevalence” (see definition in **Methods**) was 36% (95% CI: 33-40; grey column, **Figure 3A**), at which PPV and NPV were 91% (90-92) and 94% (93-94), respectively. This did not meaningfully differ with different trace recategorization strategies. From our sensitivity and specificity estimates, we estimated predictive values as a function of “prevalence”. For example, in a setting where the culture-positivity rate from a subsequently submitted specimen in patients initially culture contaminated is approximately half that observed in our cohort (vertical line, **Figure 3A**), PPV and NPV would be 80% (78-81) and 98% (97-98), respectively; with PPVs increasing to 96% (95-97) and 98% (97-98) and NPVs remaining similar at 94% (93-95) and 91% (91-92) with trace exclusion and reclassification strategies, respectively.

#### Rifampicin resistance

Predictive value as a function of the proportion of culture-contaminated patients with a later submitted TB-positive culture that was rifampicin-resistant is in **Figure 3B**. The proportion of contaminated cultures later TB-positive and rifampicin resistant was 16% (10-23), resulting in a PPV and NPV of 90% (89-91) and 99% (99-99), respectively.

#### Predictive values relative to sputum Ultra

These values were very similar to those for Ultra on sputum for TB and rifampicin susceptibility using sensitivity and specificity derived from a systematic review and meta-analysis used by the WHO for policy making (19) (**Figure 3C**).

### Potential turnaround time improvements

The potential improved TAT for TB was a median (IQR) of 42 (30-50) days overall, 23 (13-45) days for patients’ culture-positive on the repeat specimen and 49 (42-64) days for patients’ culture-negative on the repeat specimen (**Figure 2**). The improvement in TAT to MTBDR*plus-*rifampicin susceptibility testing on a positive culture isolate was 24 (15-49) days.

**Figure 2.**
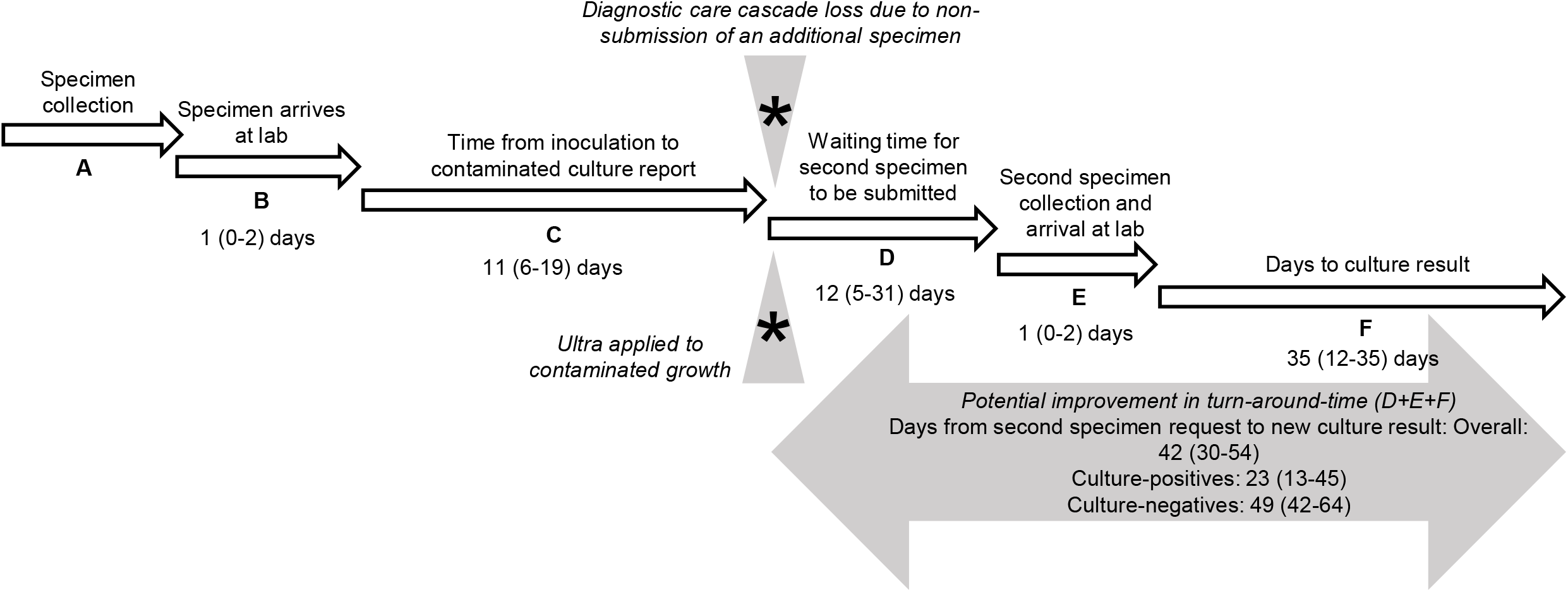
Concept map showing the timeline from. **(A)** the date of initial specimen collection to **(B)** to when it arrives at the laboratory for processing and **(C)** when it is reported as contaminated. At this point (asterisks), significant care cascade loss occurs due a subsequent specimen not being forthcoming. This loss, and the subsequent delays to await collection of another specimen **(D)**, if received at all), send it to the laboratory **(E)** and culture it **(F)** can be minimised if the Ultra on contaminated culture approach is applied. All day values are median (IQR). Abbreviations: IQR, Interquartile range; Ultra, Xpert MTB/RIF Ultra.

**Figure 3.**
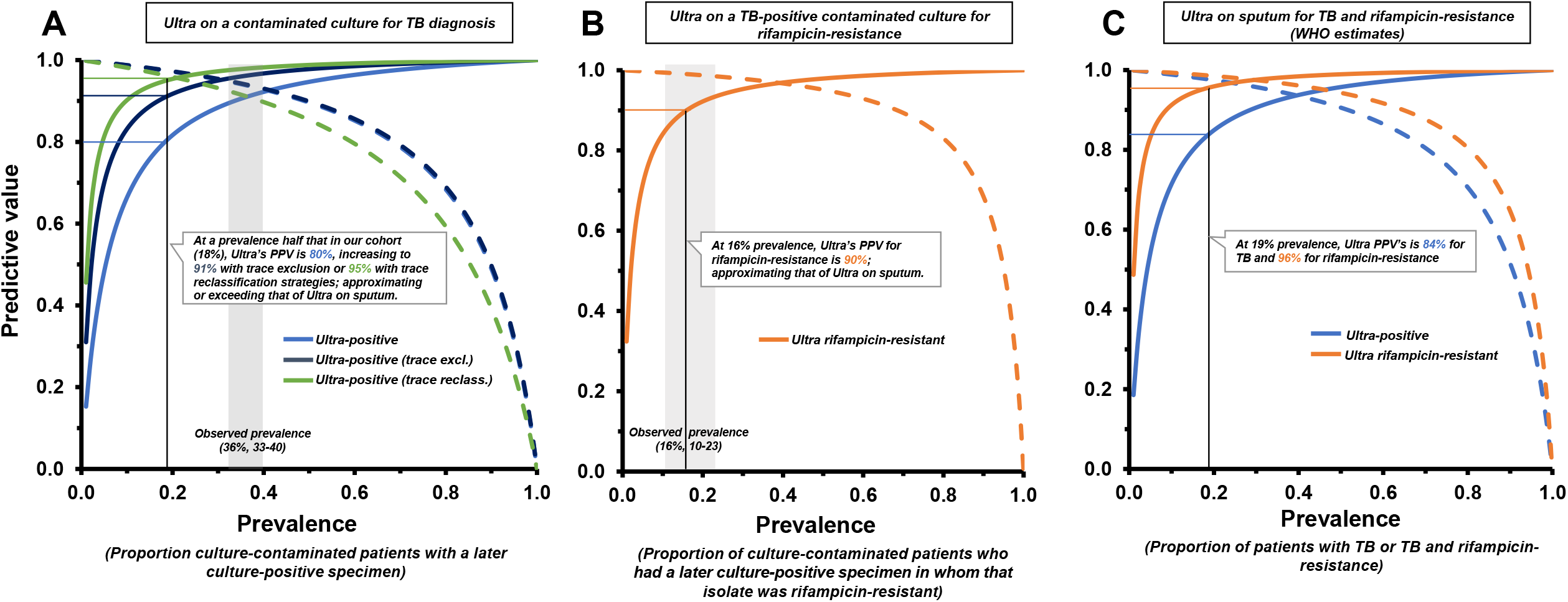
Predictive value of Ultra on AFB-negative contaminated MGIT960 growth as a function of prevalence of TB. **(A)** or rifampicin-resistance **(B)**, compared to the predictive value of Ultra on sputum per WHO estimates (19) **(C)**. Ultra on contaminated cultures approximates its predictive value on sputum for both TB and rifampicin resistance. In **(A)**, in a scenario where prevalence is half that of our cohort (vertical x-intercept line at 19%), PPV and NPV would be 81% and 97 %, respectively. PPVs increased slightly when trace recategorization strategies were used. In **(B)**, the vertical x-intercept line (16%) shows the proportion of culture-contaminated patients who had a later culture-positive specimen in whom the isolate was rifampicin resistant. At this prevalence, Ultra’s PPV and NPV on contaminated growth were 90% and 99%, respectively. Prevalence is based on results from a subsequently submitted sputum (see **Methods**). Grey columns show in-study prevalence with 95% confidence intervals. Solid lines are PPVs, dashed lines NPVs. Abbreviations: AFB, acid-fast bacilli; NPV, negative predicative value; PPV, positive predictive value; TB, tuberculosis; Ultra, Xpert MTB/RIF Ultra; WHO, World Health Organization.

### MPT64 TBc Antigen Testing

From the 20 randomly selected MGIT960 tubes where Ultra had detected TB, one (5%) was TB positive by TBc. The 20 tubes where Ultra did not detect MTBC were all TBc negative.

## Discussion

This study is the first to describe rescuing a TB result from contaminated cultures using a WHO-recommended rapid molecular test (Ultra). The key findings were: 1) Ultra on contaminated cultures is comparable to Ultra on sputum for TB detection and 2) rifampicin-resistance, and that 3) this can reduce delays associated with the need to collect and culture a second specimen but, more importantly where there is care cascade loss due to non-submission of a repeat specimen (which we show to be unfortunately frequent), it could generate a diagnostic result (with high sensitivity and specificity) where there is none. Lastly, 4) we identified many patients receiving multiple cultures simultaneously or in quick succession to one another. Together, these findings have implications for improving TB and drug-resistant TB diagnosis and can reduce the negative effects associated with liquid culture.

Our approach detected 9/10 TB cases, a sensitivity which approximates those previously reported for Ultra on sputum (2, 3). In contrast, TBc performed poorly. Furthermore, the Ultra approach had non-actionable result rates (not Ultra positive or negative) lower than those reported by other studies in our setting (3); likely because culture growth, although contaminated, is more homogenous relative to sputum. We modelled how the predictive value of our approach would change at different prevalence’s (frequency of culture-positivity in patients who had a later specimen), which creates a framework for different settings to consider rolling out our approach and showed that these predictive values mirrored those widely-accepted for Ultra on sputum. The high TB (and rifampicin resistance) prevalence’s in people who were initially culture-contaminated in our cohort was surprising, but could be biased by the types of individuals who are likely to have a repeat specimen retrieved, or due to the fact that individuals likely to be culture-contaminated have, due to TB, a perturbed respiratory microbiome (20).

Although our contaminated Ultra approach’s specificity was high, we decided to evaluate the effect of different trace recategorization strategies on sensitivity and specificity. These resulted in small specificity improvements (with a cost of excluded or missed Ultra-positives). Discordant results (Ultra-positive on the contaminated culture and subsequent culture negative) might be due to non-viable MTBC from previous TB disease as our approach had slightly diminished specificity in patients with previous TB [Xpert detects DNA from lysed cells and, as a result, patients who complete treatment can continue to be Xpert-positive for years thereafter (2, 3, 14)]. Laboratories may wish to apply the same criteria and conditions they use for reporting Ultra “trace” results on clinical specimens to “trace” results from our approach.

For rifampicin resistance, contaminated culture Ultra had high concordance with MTBDR*plus* on the repeat culture isolate. The two patients with discordant Ultra rifampicin-resistant and MTBDR*plus* rifampicin susceptible results could be due to laboratory error or heteroresistance (these individuals had no record of other specimens unfortunately, however, both were treated with the first-line regimen and died while on treatment). For the Ultra rifampicin-susceptible MTBDR*plus* rifampicin-resistant patient, a further three repeat specimens were submitted for culture and the MTBDR*plus* result was inconsistent (after the first MTBDR*plus* resistant result, the next two were MTBDR*plus* susceptible and the last was again resistant, indicating likely heteroresistance).

While our approach is likely to reduce time to diagnosis (potentially bringing a positive diagnosis forward by a month in initially culture-contaminated patients), its most significant impact is likely to occur amongst the many patients who, despite guidance to the contrary, do not have a repeat specimen submitted (almost half of individuals in our cohort). Their TB diagnoses (and potential rifampicin-resistance) is lost by the system, and we now show how this could be reduced; potentially resulting in timely TB and drug-resistance diagnoses, more effective regimens for individuals, and reduced adverse outcomes and transmission. This care cascade loss also indicates a need for algorithm strengthening in clinics (to ensure patients receive a follow-up culture). Conversely, we also observed over requesting of cultures in the laboratory, with many simultaneous or repeat cultures in rapid succession in ∼30% of patients. This is contrary to guidelines and highlights the need for laboratory strengthening, especially in terms of specimen gatekeeping systems where, for example, a culture request could be denied if another culture is underway or was shortly completed. This would result in cost savings.

A limitation of our study is that Ultra was done on a different specimen to the reference test. The time and changes in concentration of bacilli in specimens between sampling could lead to discordant results, however, our approach had excellent sensitivity and specificity. It is impossible to exclude the possibility of patients having started treatment between provision of their diagnostic specimen for culture and a repeat specimen, however, treatment would first render patients culture-negative. We would therefore expect our approach to have poor specificity but this did not occur and this scenario therefore appears unlikely. Our laboratory also uses a 35-day MGIT960 incubation period due to limited space and this may have affected sensitivity and specificity estimates. Lastly, our work should be validated in other settings, however, we have at provided proof-of-concept.

In conclusion, our results suggest that Xpert Ultra on contaminated cultures is highly accurate to establish the bacteriological diagnosis of TB. As the cost of an Ultra approximates that of culture and our approach has many potential upsides (reduced care cascade loss, improved turn-around-time), we strongly advocate for laboratories that experience contamination in TB diagnostic cultures to consider implementing our approach and assess whether this could lead to patient-level benefits.

## Supporting information

Supplementary material will be used for the link to the file on the preprint site

## Data Availability

All data produced in the present work are contained in the manuscript

## Author Contributions

Y.G., G.T. and N.B. conceptualized the experiments. Y.G., B.P., A.T., H.T., R.A., T.D. assisted with data curation. Y.G. performed formal analysis, methodology and writing original draft. G.T., N.B, assisted Y.G. with the investigation. All authors reviewed and edited the manuscript.

## Funding

RMW acknowledges funding from the South African Medical Research Council. GT acknowledges funding from the EDCTP2 programme supported by the European Union (RIA2018D-2509, PreFIT; RIA2018D-2493, SeroSelectTB; RIA2020I-3305, CAGE-TB) and the National Institutes of Health (D43TW010350; U01AI152087; U54EB027049; R01AI136894).

## Acknowledgements

The authors thank the National Health Laboratory (NHLS) as a source of data and Staff of Greenpoint Tuberculosis Laboratory, NHLS, Cape Town, South Africa for assisting with collecting contaminated MGIT960 cultures for the study.

## Notes

### Competing Interest Statement

The authors have declared no competing interest.

### Funding Statement

R.W. acknowledges funding from the South African Medical Research Council. G.T. acknowledges funding from the EDCTP2 programme supported by the European Union (RIA2018D-2509, PreFIT; RIA2018D-2493, SeroSelectTB; RIA2020I-3305, CAGE-TB) and the National Institutes of Health (D43TW010350; U01AI152087; U54EB027049; R01AI136894).

