## Supplementary material will be used for the link to the file on the preprint site for "Something from nothing: Sensitivity and specificity of Xpert MTB/RIF Ultra on contaminated liquid cultures for tuberculosis and rifampicin-resistance detection"

### Xpert MTB/RIF Ultra on AFB-negative growth from contaminated MGIT960 tubes flagged as growth positive

#### A. Purpose

This standard operating procedure (SOP) describes in detail the steps to follow when processing and performing Xpert MTB/RIF Ultra testing on Mycobacterium Growth Indicator Tube (MGIT) 960 cultures.

The workflow when growth is detected by the MGIT960 instrument is as follows:

Perform acid-fast bacilli (AFB's) smear (Ziehl Neelson or Auramine stain)
- If smear is positive for AFB's, perform standard speciation test
  - If speciation test is positive, then it is reported as positive for *Mycobacterium tuberculosis* complex (MTBC)
  - If speciation test is negative, then is reported as non-tuberculous mycobacterium (NTM)
- If smear is negative and contaminating organisms are observed, then perform Xpert MTB/RIF (Ultra)
  - If Ultra is positive, then report as positive for MTBC
  - If Ultra is negative, then report as negative for MTBC

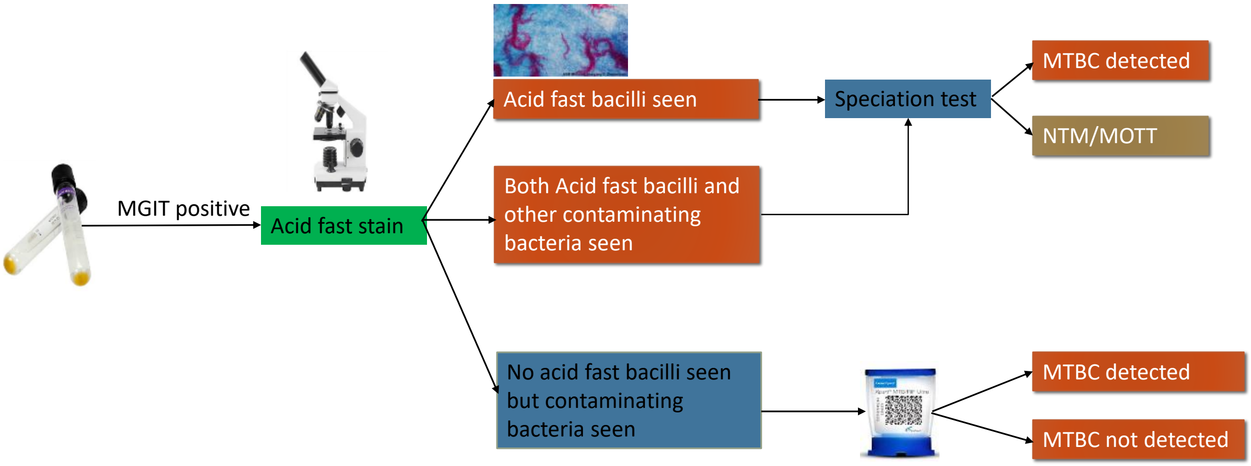


#### B. Supplies needed

1. Biosafety Cabinet
2. Xpert Ultra kit (Xpert Ultra cartridges, Sample Reagents (SR) and sterile transfer pipettes) Xpert DX system (GeneXpert instrument, computer, software, scanner, printer, and UPS) 5 ml centrifuge tubes
3. Sterile >15ml centrifuge tubes
4. Centrifuge
5. Pasteur pipettes P1000 pipette filter tips
6. Timer (stopwatch)

**Notes:**
- Store the Xpert Ultra cartridges and reagents at 2 to 28 °C
- Do not use cartridges or reagents beyond the expiration date
- Do not open cartridge lid until ready to perform the testing

#### C. Responsibilities

1. All staff working in the Laboratory are responsible for the implementation of this Standard operating procedure.
2. All users of this procedure who do not understand it or are unable to carry it out as described are responsible for seeking advice from their supervisor.
3. It is the responsibility of the lab personnel to follow this SOP as it is written.
4. It is the responsibility of the lab personnel to follow the necessary safety precautions when handling MGIT960 cultures during this process.

#### D. Safety

1. Only trained personnel will perform Ultra testing in MGIT960 contaminated cultures
2. Before you start to use the Gene Xpert Dx system, make sure you read the operator manual available in the lab. Using controls, making adjustments, or performing procedures other than those specified in this SOP may result in hazard or injury to person or damage to the equipment.
3. Do not attempt to lift the instrument without proper safety training and assistance. This may damage the instrument and void the warranty.
4. Read and follow the instructions in the product insert of Ultra cartridge.
5. All processing of contaminated MGIT960 cultures should be performed as per the biosafety standards.
6. Treat all MGIT960 cultures, including used cartridges, as potentially infectious.
7. Wear protective gloves and laboratory coats when handling specimens and reagents.
8. Wash hands thoroughly with soap and water after handling specimens and test reagents.
9. All benches should be decontaminated after work, or immediately after a spill, with an appropriate Mycobactericidal disinfectant, followed by 70% alcohol.
10. Do not reuse Ultra cartridges.
11. Dispose of used Ultra cartridges according to infectious waste material disposal guidelines.

#### E. Processing and testing.

1. A MGIT tube containing 7 to 8 ml of liquid culture will be tested using the Ultra assay.
2. If the volume is >7ml; 6 ml will be centrifuged at 3000 xg for 15 minutes and supernatant will be discarded into a waste container with appropriate disinfectant. The pellet will be used for further Ultra testing.
3. If the volume is <7 ml; 1 ml of the culture will be stored, and the leftover will be centrifuged at 3000 xg for 15 minutes and supernatant will be discarded into a waste container with appropriate disinfectant. The pellet will be used for further Ultra testing.
4. Any leftover culture will be stored at -20°C for further investigation if needed.

#### F. If Volume of MGIT is ≥ 7 ml:

1. Label one 50/15 ml centrifuge tube with the patient study number, date and specimen type i.e. “MGIT”.
2. Transfer 6 ml of the culture to the centrifuge tube and concentrate by spinning at 3000 xg for 15 minutes at room temperature. Store the rest of the culture in the MGIT960 tube.
3. Using a sterile Pasteur pipette, remove the supernatant and discard into appropriate disinfectant, leaving a volume of 0.7ml of the supernatant and vortex thoroughly.
4. Add 1.4 ml of Ultra sample reagent buffer using a Pasteur pipette. The treated specimen can only be used until 4 hours from the addition of sample reagent.
5. Seal the lid of the centrifuge tube tightly and vortex thoroughly for 60 seconds and until no clumps are observed.
6. Incubate for 10 minutes at room temperature
7. Vortex thoroughly for 60 seconds.

8. Incubate the sample at room temperature for an additional 5 minutes
9. Label the Ultra cartridge with the sample study number
10. Mix sample by swirling gently
11. Using a sterile Pasteur pipette, transfer 2 ml of the sample and reagent mixture to the Ultra cartridge, avoiding bubbles
12. Load cartridge into the GeneXpert instrument as per manufacturer's instructions as soon as possible.

**G. If Volume of MGIT is < 7 ml**

1. Label a 50 ml centrifuge tube with the patient study number, date and specimen type i.e. "MGIT"
2. Store 1 ml of the MGIT culture in a microcentrifuge tube and transfer the remaining culture to the 50 ml tube
3. Concentrate culture by spinning at 3000 x g for 15 minutes at room temperature

**H. Repeating Ultra using unconcentrated MGIT:**

1. This is to be used in the event of Ultra test resulting in non-actionable result that would require the Ultra to be performed again.

2. Non-actionable result occurs when the assay run is unsuccessful and is reported as an "invalid, no-results or error" by the GeneXpert instrument. This could be due to system control failure, improper sample preparation, power interruption or failure etc.

    A. Add 2 ml of Ultra sample reagent buffer to the 1 ml aliquot of MGIT culture
    B. Vortex thoroughly for 60 seconds
    C. Incubate at room temperature for 10 minutes
    D. Vortex thoroughly for 60 seconds
    E. Incubate at room temperature for a further 5 minutes
    F. Add 2 ml sample- Xpert reagent mixture into a correctly labelled Ultra cartridge, avoiding bubbles
    G. Load cartridge into the Xpert instrument as per manufacturer's instructions
